# Global governance of pandemic prevention from the wildlife trade: A perspective from governance entrepreneurs and practitioners

**DOI:** 10.64898/2026.07.08.26357525

**Authors:** Ryan Gray, Eduardo Gallo-Cajiao, Raphael Aguiar, Kirsten M. Lee, Tarra L. Penney, Mary Wiktorowicz

**Author notes:** Corresponding author: (EGC).

## Abstract

Although a strand of scholarship on pandemic prevention flourished in the wake of the COVID-19 pandemic, a theoretically informed empirical analysis of global governance entrepreneurs and practitioner perspectives is lacking. This gap is salient given the need to consider the nuances, political realities, and feasibility of real-world governance practice, particularly with the recent adoption of the Pandemic Agreement under the World Health Organisation. In this paper, nexus governance and regime complex theory guides an analysis of recommendations for potential real-world governance responses for pandemic prevention from wildlife trade for human consumption elicited from global governance entrepreneurs and practitioners through semi-structured interviews and document analysis. Recommendations on future governance practice largely focused on strengthening coordination across various policy sectors to improve use of existing institutional arrangements, with particular emphasis on better integration of the biodiversity conservation policy sector within global pandemic prevention governance, as well as reform of the World Organisation for Animal Health and the Convention on International Trade in Endangered Species of Fauna and Flora. With governance deficits for prevention of pandemics emerging from the wildlife trade left by the now largely concluded Pandemic Agreement, a renewed research agenda on shared governance pathways becomes paramount.

## Introduction

The SARS-CoV-2 (COVID-19) pandemic revealed serious deficits in global governance for pandemic prevention [1], making evident the need for a shift in policy paradigm from downstream to upstream approaches. On one hand, there were shortcomings in the implementation of the International Health Regulations (IHR) [2,3], the main governance architecture for global pandemic prevention available at the time [4]. On the other hand, COVID-19 revealed a lack of governance mechanisms for upstream prevention by focusing on surveillance, early detection and containment in human populations [5]. Governance of upstream prevention of zoonotic spillover events [6] is a priority given the role of the wildlife trade and hypermobile transportation networks that hasten emerging infectious disease spread [7–9].

Against this backdrop, an Agreement on Pandemic Prevention, Preparedness, and Response (Pandemic Agreement hereafter) under the World Health Organisation (WHO) was adopted in 2025 [10] as a central tool for addressing the challenges and gaps within global health governance [11]. However, the Pandemic Agreement did not realise its full potential, critiqued by policymakers and scholars for its lack of sufficient accountability, obligations weakened by sovereignty concerns, and the absence of key actors from negotiations and adoption, including multilateral stakeholders and influential states [12–16]. Although the Pandemic Agreement calls for Parties to take measures to prevent zoonotic spillover, it does not adequately address operationalisable strategies to that end [17], warranting further scholarly attention as to how governance of pandemic prevention can be strengthened into the future.

While global health governance scholarship has begun to inform governance responses for the prevention of future pandemics resulting from zoonotic spillover, a theoretically guided empirical analysis of potential solutions from governance entrepreneurs and practitioners is generally lacking. Where the perspectives of practitioners are considered [18–21], they apply to a specific policy sector, issue, and geographic area rather than generally to the governance of global pandemic prevention. This shortcoming is particularly salient given that the governance response for pandemic prevention in relation to the wildlife trade for human consumption (wildlife trade hereafter) involves human and animal health, biodiversity conservation, trade and food governance systems, with actors spanning multiple policy sectors, governance levels, geographic scopes, and sectors of society [22–24]. Addressing this lacuna can provide pragmatic insights to contribute to governance responses and scholarship addressing spillover risks, as governance entrepreneurs and practitioners are well positioned to understand the feasibility and limitations of formulating and implementing governance solutions [25].

Against this backdrop, here we present, compile, and analyse potential upstream pandemic prevention governance responses addressing the risks of zoonotic spillover from the wildlife trade. Through a thematic analysis of semi-structured interviews of governance entrepreneurs and practitioners, as well as documents referred to during interviews, we specifically ask: (1) what are the constraints and alternatives for institutional responses banning wildlife trade to foster pandemic prevention from the wildlife trade for human consumption?; (2) what are the constraints and opportunities to better use existing institutional arrangements to foster pandemic prevention from the wildlife trade for human consumption?; (3) what are the constraints and opportunities for institutional reform to foster pandemic prevention from the wildlife trade for human consumption?; and (4) what are the constraints and opportunities for institution building to foster pandemic prevention from the wildlife trade for human consumption? This theoretically guided empirical analysis adopts a global scope and contributes to the emerging scholarship on pandemic prevention governance, focusing on pragmatic approaches with potential for implementation.

### Theoretical framework

Within global health governance, pandemic prevention requires cross-sectoral and multi-level coordination that is made difficult by issues of institutional fragmentation, multiplicity, and gridlock [26,27]. In the current polycrisis, the capacity of individual institutional arrangements for problem solving becomes outpaced by emerging challenges. Institutions that primarily focus on single problems struggle to coordinate with other single-issue institutions, and limited understanding exists of the drivers of global crises and the impact of system-wide interactions [28]. Recent developments in global health governance attempt to foster cross-sectoral collaboration and integration through One Health [29]. One Health recognises that human health, animal health, environmental health, and food security are interdependent [30] and aims to address global health issues through better coordination, collaboration, and communication between policy sectors. However, implementation challenges occur at the state level, including conflicting and unclear values and priorities, limited political will and leadership, insufficient funding and human resources, limited coordination, and inadequate surveillance and data sharing across policy sectors [31].

In this analysis, we conceptualise the global governance system for pandemic prevention through a nexus governance approach and as a regime complex. Nexus governance focuses on the dismantling of policy silos and the integration of governance systems and actors to maximise synergies and co-benefits, minimise trade-offs, and enhance sustainability, originally developed for management of the water, energy, and food nexus [32]. The governance of pandemic prevention is highly multisectoral, consisting of global health governance, global biodiversity conservation governance, global food governance, and global trade governance [24], thus requiring the coordination of distinct governance systems. Regime complexes are defined as a group of overlapping and non-hierarchical international institutions, agreements, principles, norms, and rules that govern a particular issue area [33]. As the product of multiple governance systems, global pandemic prevention governance has been characterised as highly fragmented, containing overlapping regulatory institutional arrangements applied in a patchwork manner [22] that lacks a supranational body to coordinate and adjudicate conflicts, overlaps, and inconsistencies between institutional arrangements. The regime complex approach highlights power relations and dynamics among actors, the effectiveness of policy outcomes, and conditions under which regime complexes hinder or facilitate cooperation [33]. Our analysis aims to highlight the relational structures and processes that shape global governance processes and outcomes, connect actors, and inform discourse [34]. Thus, we conceptualise global health governance as the governance arrangements needed to advance global health goals and principles [35], and in the case of nexus governance and regime complexes, how to best manage institutional fragmentation, multiplicity, and overlap through coordinated governance architectures.

##### Text Box 1. Definition of key concepts

###### Global governance

The overarching system of state and non-state actors, institutions, and organisations, principles, norms, regulations, and decision-making procedures that are active in a given issue area of global politics [36]. Global governance is characterised by the absence of a central supranational authority, the emergence of non-state actors, the need to adapt to and manage the externalities arising from increased transboundary flows, and the consideration of scales from the local to the supranational [35].

###### Institutional arrangement

The formal and informal structures, rules, norms, systems, practices, and processes, that are mutually agreed upon and that are paper based, and which guide how actors plan, coordinate, implement, and monitor activities to achieve goals and define roles, responsibilities, and relationships between such.

###### Wildlife trade

The trade of any organism, including fungi, plants and animals, sourced from the wild, regardless of geographic scope or scale [37].

###### Zoonotic spillover

The process that enables a pathogen from a vertebrate animal to establish infection in a human [38].

###### Fragmentation

The diffusion of responsibility across multiple actors within the global health governance system as a result of the exponential increase in actors engaging within the system, resulting in overlapping mandates, duplicated efforts, increased transaction costs, greater competition for resources, and the influence of powerful countries and actors on the global health agenda [27].

###### Sectoral fragmentation

The diffusion of responsibility within the global health governance system across multiple policy sectors because of growing demand for organisational specialisation, uniqueness, and functional differentiation, resulting in the siloisation of policy sectors that prioritise sectoral agendas rather than cohesive, system-wide agendas [39].

###### Policy coherence

The systematic promotion of mutually reinforcing prescriptions across policy sectors at all geographic scopes, creating synergies towards achieving a mutually agreed vision or objectives so that efforts in one policy sector do not undermine efforts in another while reinforcing those efforts where possible [40].

###### Policy incoherence

Policy actions, objectives, or outcomes contradicting, undermining, or impeding one another because of actors failing to align objectives and policy actions across policy sectors.

## Methods

### Study design and data collection

This study consisted of a single case study of semi-structured interviews with globally based key informants (KIs), conducted from June 2021 to September 2022 by MW and EGC. The study was approved by the Human Participants Review Sub-Committee of York University’s Ethics Review Board (certificate #e2020-310) and conforms to the standards of the Canadian Tri-Council Research Ethics guidelines. This timeframe coincides with the unfolding of the COVID-19 pandemic. Informants were either governance entrepreneurs or practitioners involved in issues related to the governance of the wildlife trade, human health, animal health, international trade, food security, and the prevention of emerging zoonoses (Table 1). Global governance entrepreneurs are individuals who are not necessarily based in intergovernmental organisations, treaty secretariats, or nongovernmental organisations, but who influence or transform governance structures or processes to drive or achieve goals within an institution or policy sector [41].

**Table 1.**
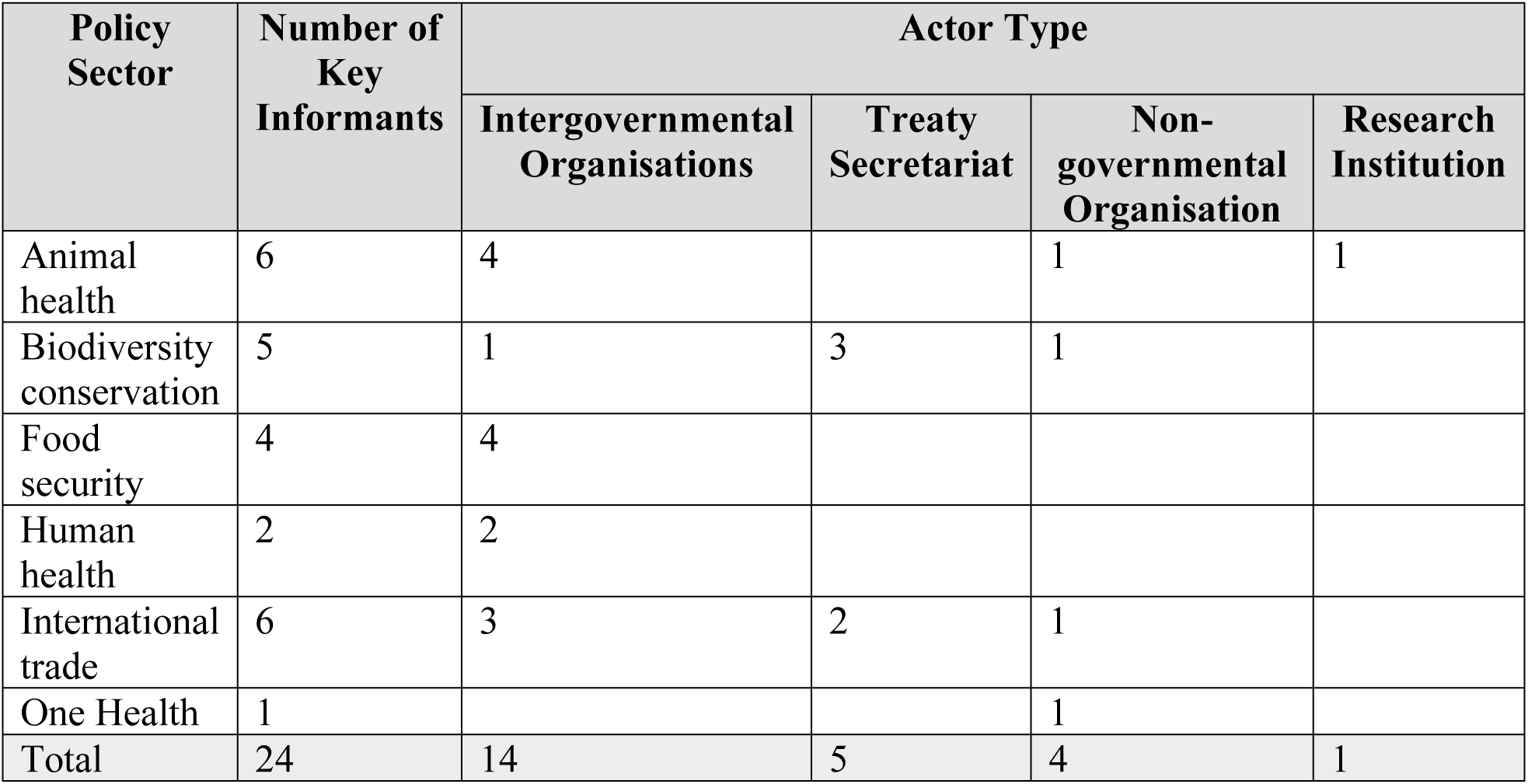
Composition of policy sector representation by number of key informant and actor type.

| Policy Sector | Number of Key Informants | Actor Type |  |  |  |
| --- | --- | --- | --- | --- | --- |
|  |  | Intergovernmental Organisations | Treaty Secretariat | Non-governmental Organisation | Research Institution |
| Animal health | 6 | 4 |  | 1 | 1 |
| Biodiversity conservation | 5 | 1 | 3 | 1 |  |
| Food security | 4 | 4 |  |  |  |
| Human health | 2 | 2 |  |  |  |
| International trade | 6 | 3 | 2 | 1 |  |
| One Health | 1 |  |  | 1 |  |
| Total | 24 | 14 | 5 | 4 | 1 |

Global governance practitioners are policymakers or executive staff based in intergovernmental organisations, treaty secretariats, or nongovernmental organisations who lead or support institutional decision or policy making. Recruitment of key informants began purposively with a seed sample by identifying participants via professional networks and desktop searches for relevant organisations based on relevant expertise and alignment with our research aims, supplemented by snowball sampling. Participants were recruited until no new relevant knowledge was elicited during interviews and thematic saturation was reached [42]. KI recruitment and data collection began on June 1, 2021 and ended on September 15, 2022. One KI provided verbal informed consent before their interview, witnessed by MW and EGC. The remaining 22 KIs signed written consent forms. Each interview lasted approximately an hour and was conducted through the online video platform *Zoom*. The interview guide focused on topics including international wildlife trade management; response to zoonotic disease outbreaks and pandemics; knowledge and causality; and coordination in efforts to govern the wildlife trade and reduce related public health risks (S1 Text). Additionally, we retrieved documents referred to by KIs as part of their answers to interview guide questions. The analysis focused on KI quotes that involved recommendations.

### Data analysis

Recommendations encompass normative or empirical statements regarding what future pandemic prevention governance in relation to the wildlife trade should, could, or will entail. Quotes were deemed to involve recommendations if they contained reference to keywords that denote normative or empirical recommendations, such as ‘need’, ‘want’, ‘have’, ‘would’, and ‘should’. As this process was iterative, relevant keywords may not be present in all sentences within a quote and instead may be contained within the context, elaboration, and/or the core sentence(s) or may be implicit. In these cases, quotes were included to ensure potentially relevant data was not prematurely excluded before further consideration for relevance. We adopted both a realist and a phenomenological approach [43,44], whereby statements from KIs represent reality, such as the existence of institutional arrangements, as well as insights anchored in lived experiences, such as perceptions of institutional performance.

Interview transcripts were read through once by one author (RG) to gather quotes from KIs concerning recommendations for the global governance of pandemic prevention. A total of 255 quotes were initially selected from the interview transcripts for further consideration. These selected quotes from the interview transcripts underwent one round of screening by one author (RG) based on the inclusion/exclusion criteria (Table 2). Finally, 44 quotes from the interview transcripts were included and 211 quotes were excluded from the analysis (Fig 1).

**Fig 1.**
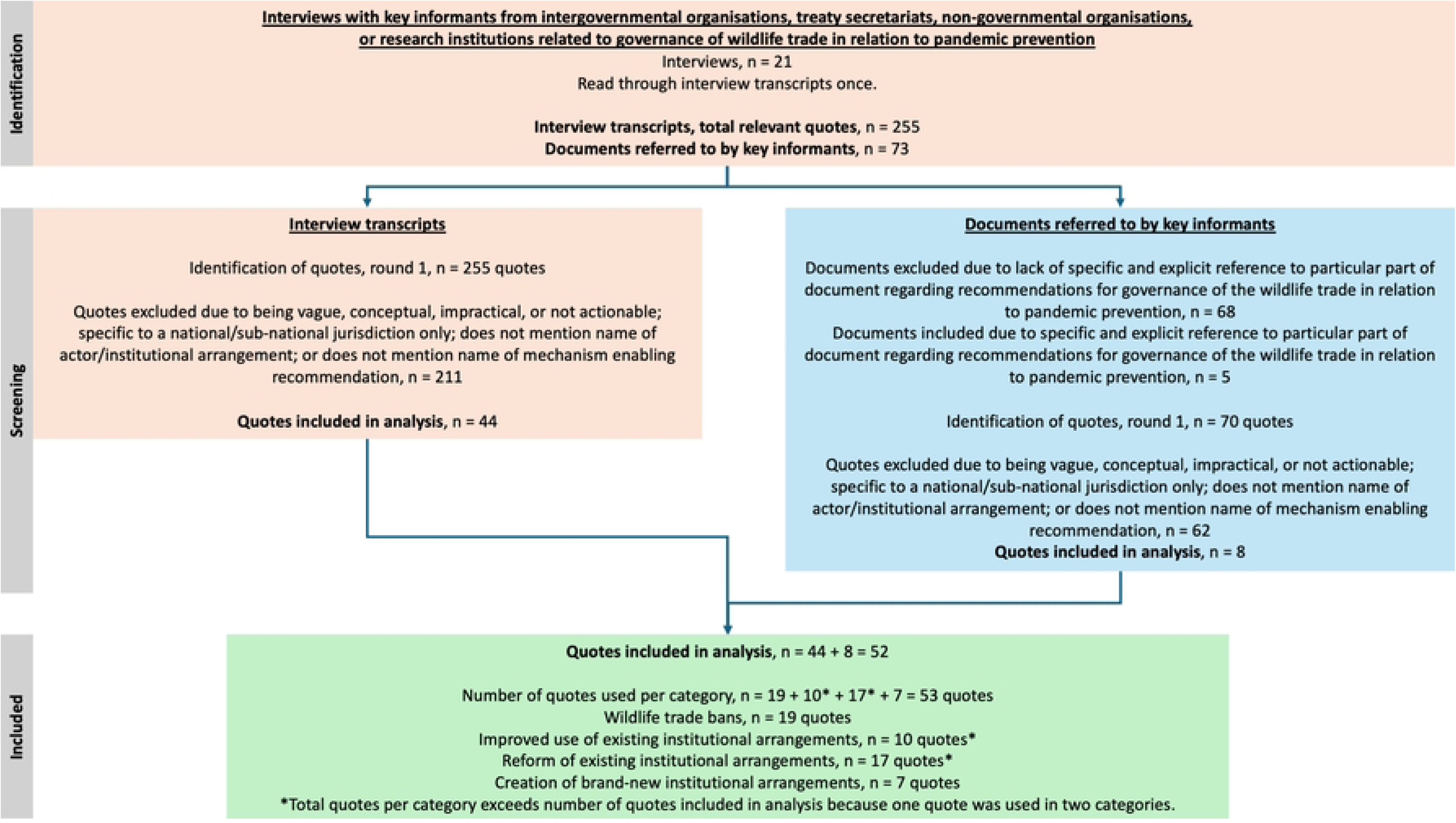
Data identification flow diagram. Flow diagram for the identification, screening, and selection of quotes from data sources.

**Table 2.**
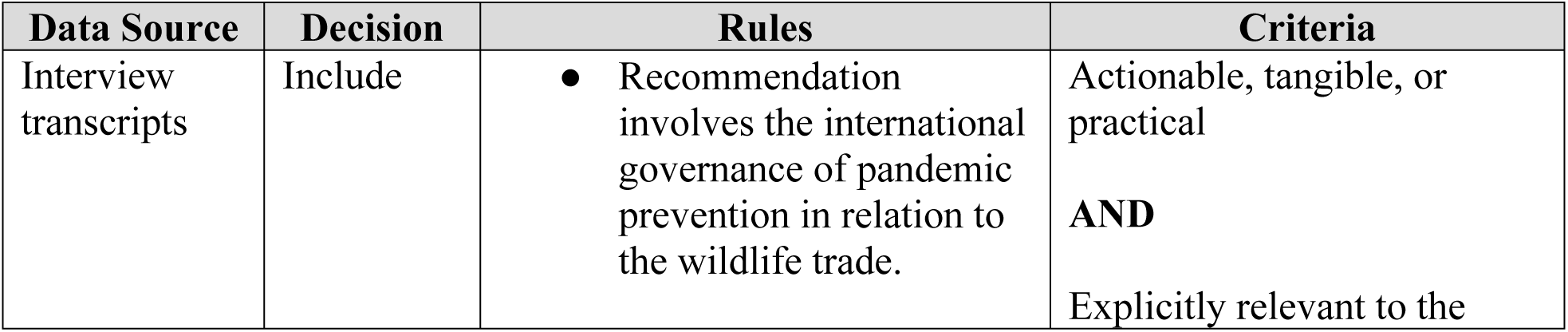
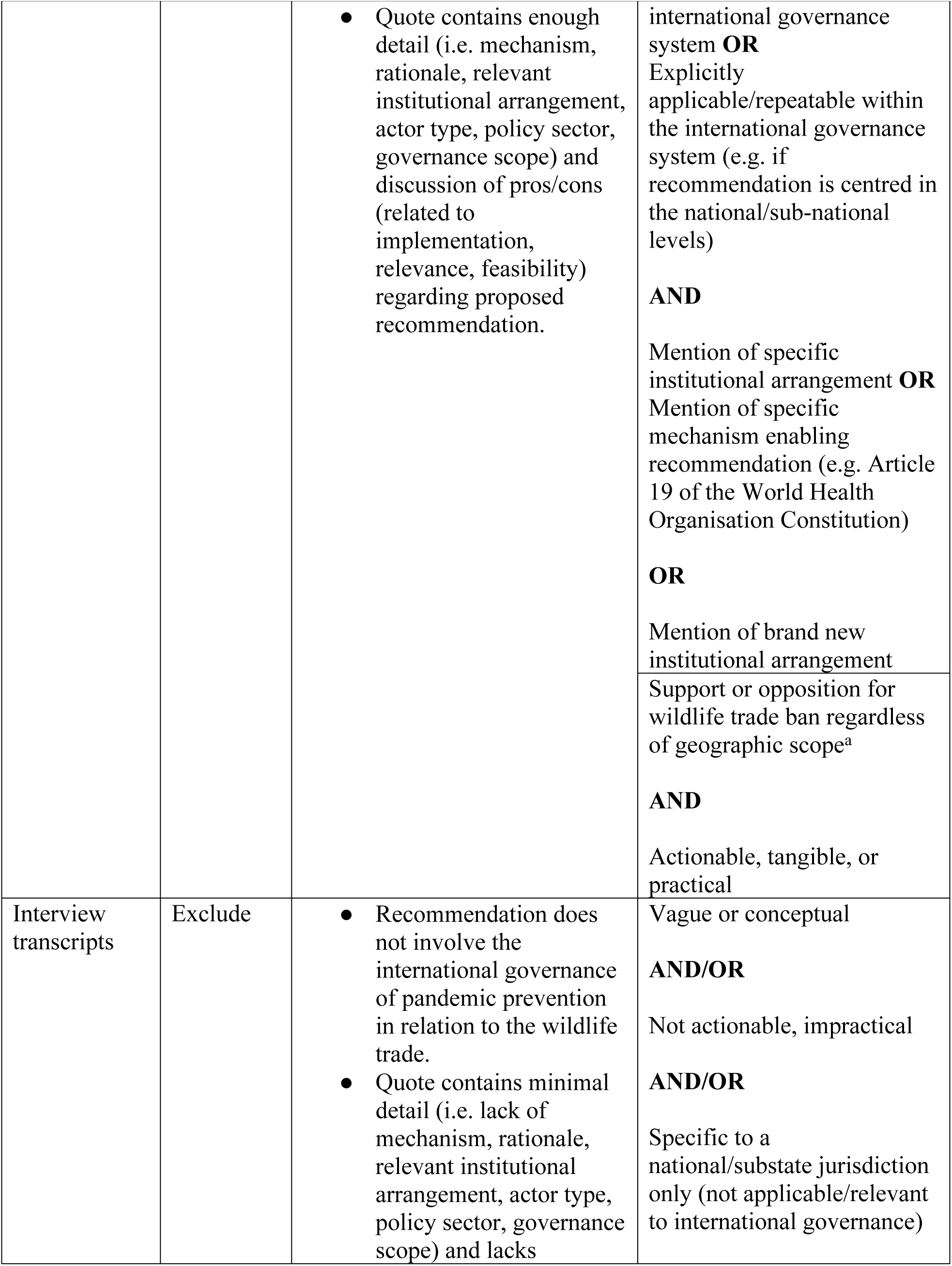
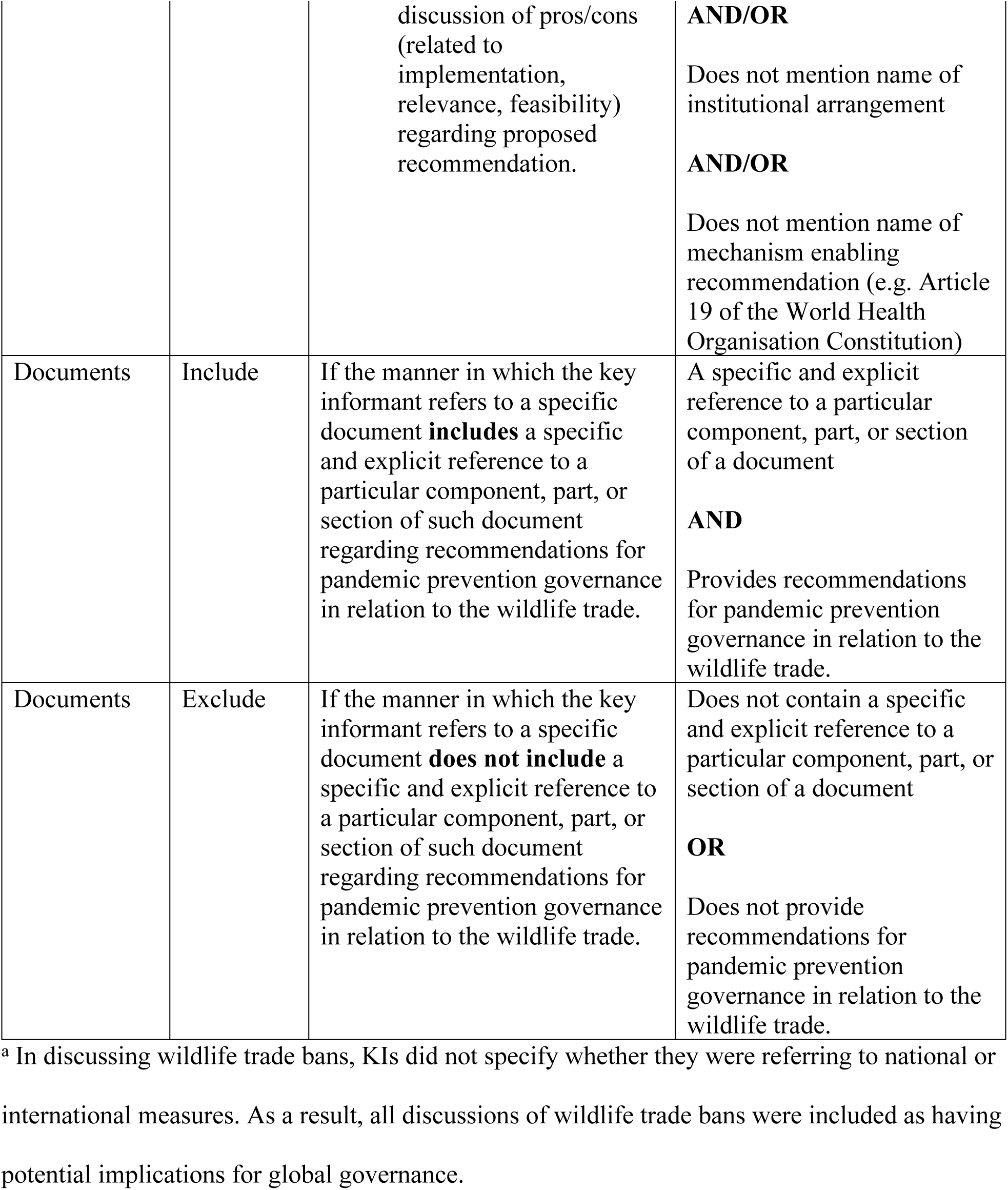
Inclusion/exclusion criteria for quotes from interview transcripts and documents.

Similarly, KIs referred to specific documents providing further elaboration when answering interview guide questions. Of 73 documents initially referred to by KIs in interviews, five documents were included and 68 documents were excluded from the analysis after one author (RG) reviewed the context of document mentions within the quotes. All excluded documents failed to refer to a particular part or section of the document (Table 2). The specific sections of the documents referred to by KIs were read through and quotes were collected by one author (RG) if they involved recommendations. Selected quotes from the documents underwent one round of inclusion/exclusion by one author (RG) based on the inclusion/exclusion criteria (Table 2). From the documents included, eight quotes were included into and 62 were excluded from the analysis (Fig 1).

The inclusion and exclusion processes for quotes from interview transcripts and excerpts from documents were intended to identify quotes that contained sufficient specific detail to allow for potential real-world application. This first phase was as inclusive as possible to ensure that any potentially relevant data was examined by the authors before being excluded.

#### Qualitative analysis

Inductive and deductive thematic analysis of semi-structured interview transcripts and documents referred to by KIs was conducted using the collaborative mixed methods platform *Dedoose* [45]. This hybrid approach combining inductive and deductive approaches has been similarly taken [46,47]. A preliminary codebook (S2 Table) was created and defined deductively by three authors (EGC, MW, RG) based on a scoping review on the regulations governing the wildlife trade in relation to zoonotic spillover prevention [22] as it mapped key concepts around regulations, gaps, challenges, and implications. The codebook includes codes that categorises the recommendations raised by KIs to appropriate discursive policy positions and is organised by drawing on nexus governance and regime complex theory, such that the codes could be applied to any recommendation regardless of its policy position to thematise the recommendations raised by KIs. Practice coding sessions were conducted by three authors (RG, EGC, MW) to further clarify code definitions and resolve ambiguities in code application.

Included quotes were iteratively categorised into one of the four following policy positions based on keywords (Table 3): 1) wildlife trade bans; 2) improved use of existing institutional arrangements; 3) reform of existing institutional arrangements; and 4) brand-new institutional arrangements. The policy positions emerged inductively from the data through the coding process and discussion between authors (EGC, RG). Finally, code applications were iteratively categorised as an opportunity, constraint, or alternative to implementing the recommendation within the respective quote.

**Table 3.** Definitions of the discursive policy positions and their categories.

| **Policy Position** | **Definition** | **Example Keywords** |
| --- | --- | --- |
| **Wildlife trade bans** | The prohibition of the legal commercialised trade of wild animals and their parts, without distinction of jurisdiction (i.e. sub-national, national, regional, international), type of wildlife (i.e. specific species or complete taxonomic groups), and purpose of wildlife use (e.g. bans of some selected uses or complete bans on all uses) [37]. | Ban; blanket; subset |
| Alternatives | Code applications that denote recommendations to be explicitly pursued in lieu of wildlife trade bans. | Need; instead |
| Constraints | Code applications that denote the lack of potential for the real-world implementation of a recommendation given the explicit mention of barriers within the quote (e.g. knowledge needs or uncertainty prohibiting future action or a lack of feasibility due to power dynamics or insufficient funding or human resources) or rationales against enacting bans. | Underground economy; human rights; unintended consequences |
| **Improved use of existing institutional arrangements** | Increased awareness of existing institutional arrangements and their potential applicability and potential changes in the relationships between existing institutional relationships. | Coordination; collaboration; make use; existing; relationship |
| Opportunities | Code applications that denote the potential for the real-world implementation of a recommendation given the explicit mention of favourable conditions within the quote (e.g. providing a precedent or denoting feasibility) or the lack of explicit mention of constraints. | Complement; working group; joint committee |
| Constraints | Code applications that denote the lack of potential for the real-world implementation of a recommendation given the explicit mention of barriers within the quote (e.g. knowledge needs or uncertainty prohibiting future action or a lack of feasibility due to power dynamics, insufficient funding or human resources). | Compete; conflict |
| **Reform of existing institutional arrangements** | The modification or expansion of the structure, function, mandate, membership, or prescriptions of already existing institutional arrangements to improve the intrinsic effectiveness or appropriateness of a given institutional arrangement. | Reform; change; expand; mandate; make use; existing |
| Opportunities | See Opportunities above. | Expand; join; add |
| Constraints | See Constraints above. | Dilute; weaken |
| **Creation of brand-new institutional arrangements** | The creation of a new global institutional arrangement for pandemic prevention that either replaces an existing institutional arrangement [48,49] or exists in tandem with other extant institutional arrangements [50]. | New; institution; actor; create; build; establish; develop |
| Opportunities | See Opportunities above. | Commit; implement; deliver on |
| Constraints | See Constraints above. | Interfere; tension |

Despite partial or complete (i.e. blanket) bans on the wildlife trade being closer to policy responses [51] rather than arrangements of institutional arrangements as the other discursive positions focus on, wildlife trade bans are nevertheless conceived as a governance response. This perspective is based on a broadened definition of institutional arrangements which highlights the rules and practices that institutional arrangements use to make decisions and exercise authority. While wildlife trade bans have not been enacted internationally, bans may materialise as a product of international agreements, for example. This precedent is demonstrated through international agreements in climate change (e.g. the Montreal Protocol on Substances that Deplete the Ozone Layer [52]; and a Global Plastic Pollution Treaty currently being negotiated through the United Nations Environment Programme [53]), and weapons regulation (e.g. the Convention on the Prohibition of the Use, Stockpiling, Production and Transfer of Anti-Personnel Mines and on their Destruction [54]; the Treaty on the Prohibition of Nuclear Weapons [55]; and the Convention on Cluster Munitions [56]) that expressly or effectively ban certain goods or practices deemed harmful. As a governance response, wildlife trade bans consist of constellations or arrangements of supporters and opposers who differ in actor type, policy sector, governance level, and geographic boundary, enabling comparison to the other discursive policy positions. Furthermore, wildlife trade bans were used to approximate zoonotic spillover emerging specifically from the wildlife trade as opposed to other pathways.

The included quotes from the interview transcripts and the documents were coded by one author (RG). The codebook was reflexively edited to allow for the addition of relevant categories and codes, and refinement of code definitions for improved application. The code applications were validated by two authors (EGC, RG) until consensus was reached.

We provide the results from referred documents within S3 Text as this information is publicly available as opposed to the results from the interview transcripts which are not publicly available due to privacy and ethical restrictions. In quantitative analyses and data visualisations, we include and consider results from both sources equally.

#### Quantitative analysis

Quantitative analyses (i.e. descriptive statistics, network graph, and degree centrality) were conducted to summarise code application, highlight patterns, reveal and highlight other prominent KI recommendations related to the research questions, and enable greater abstraction of the recommendations in order to increase broader applicability beyond specific prescriptions. The extent to which KIs discuss specific recommendations is demonstrated by the frequency of code applications that categorise quotes into discursive categories. Bar graphs visualising this frequency of code applications were plotted to determine which discursive categories and specific recommendations were most frequent among KIs (Figs 2 & 3; S4 Table).

**Fig 2.**
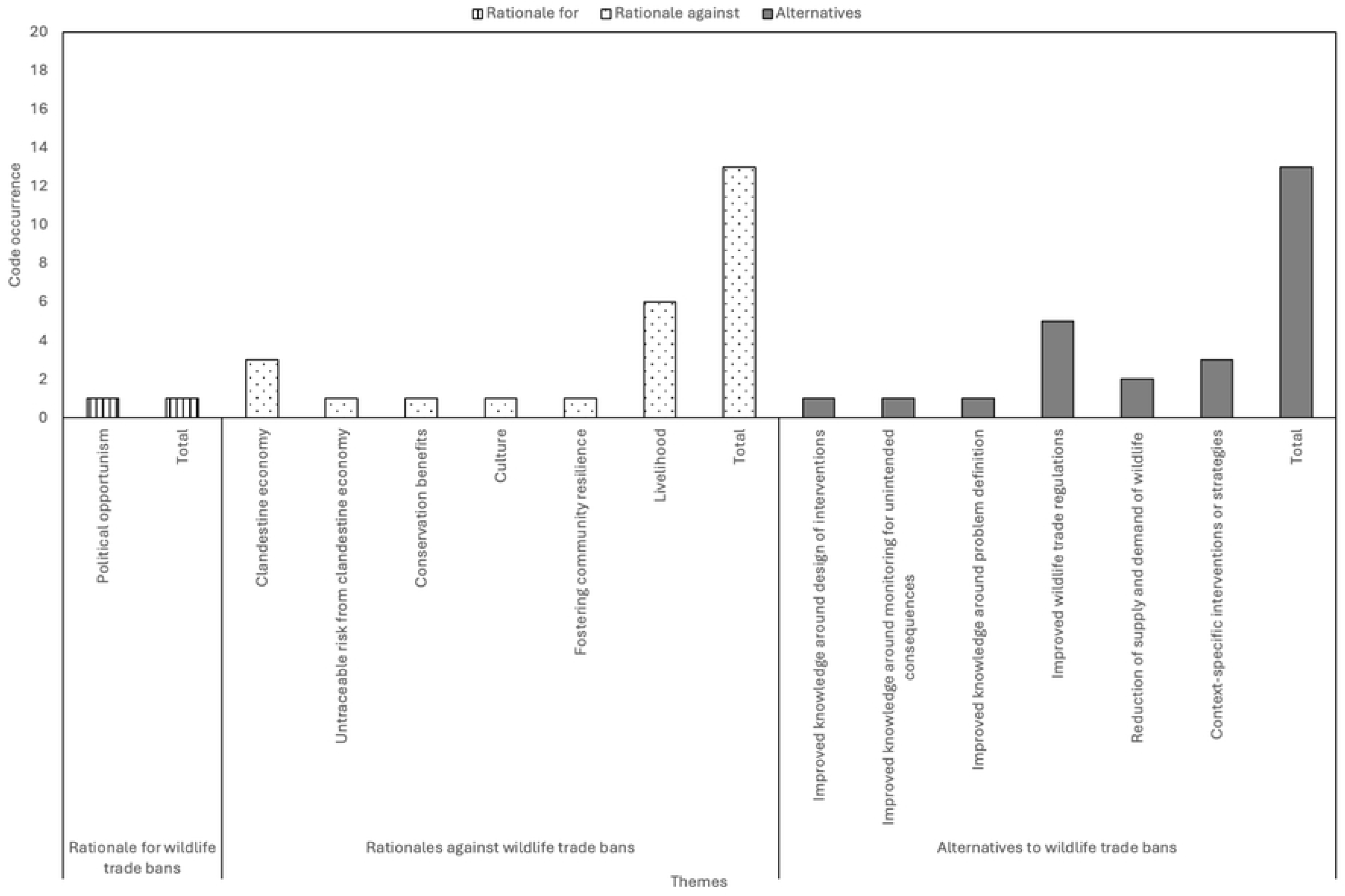
Bar chart for wildlife trade discourse. Frequency of occurrence of wildlife trade discourse by key informant, categorised by opportunity and constraint.

**Fig 3.**
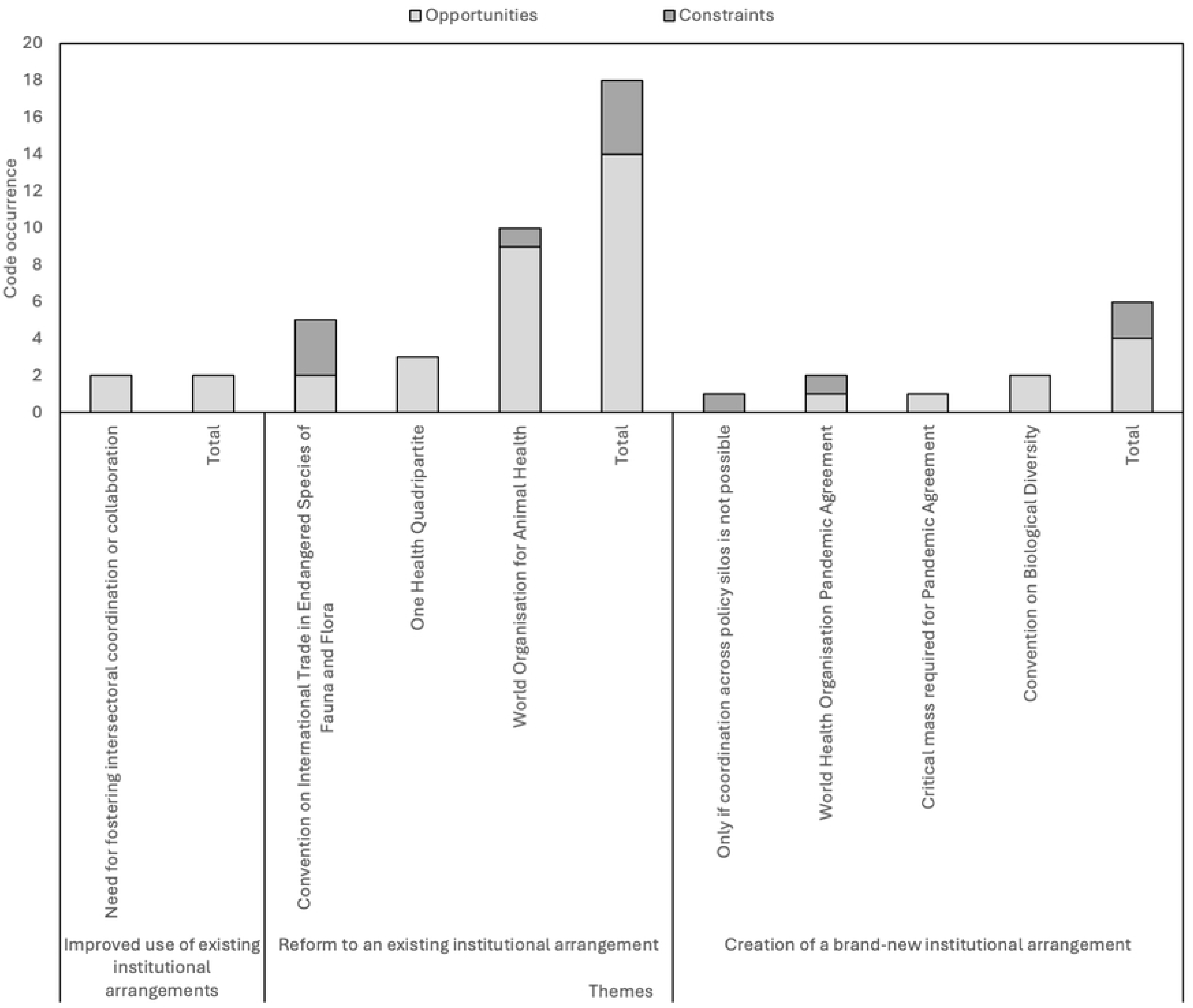
Bar chart for institutional arrangement discourse. Frequency of occurrence of improved use of existing institutional arrangements, reform of existing institutional arrangements, and creation of brand-new institutional arrangements discourse by key informant, categorised by constraint and alternative.

The categorisation of specific recommendations into opportunities, constraints, alternatives, theories, and categories is demonstrated through Alluvial diagrams (Figs 4-7) and are organized by each policy position. Alluvial diagrams were created using data visualisation software *Flourish* [58]. Corresponding tables used to plot the diagrams are contained in S5-8 Tables.

**Fig 4.**
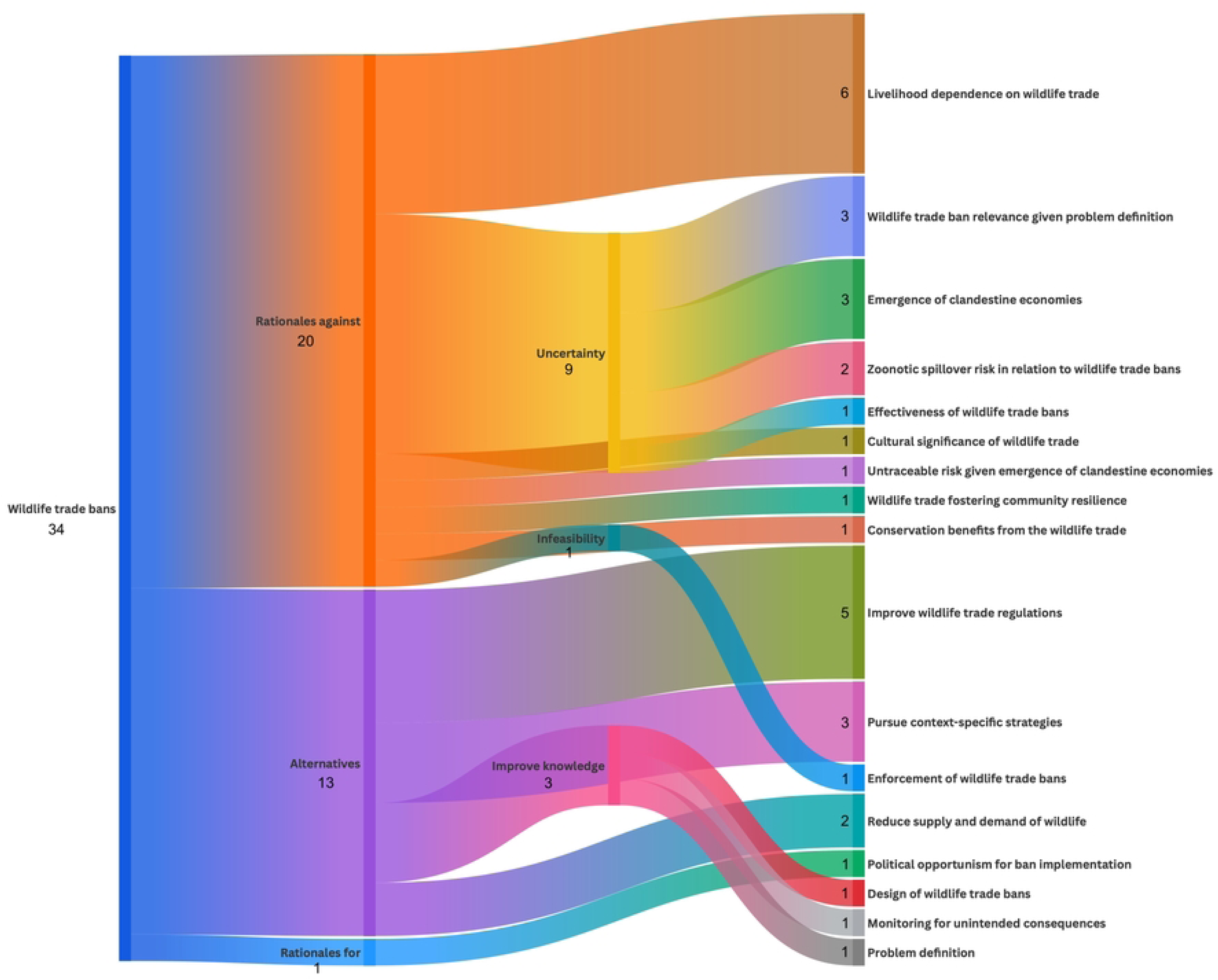
Alluvial diagram for wildlife trade bans. Alluvial diagram illustrating flows of opportunities and constraints in response to recommendations for wildlife trade bans.

**Fig 5.**
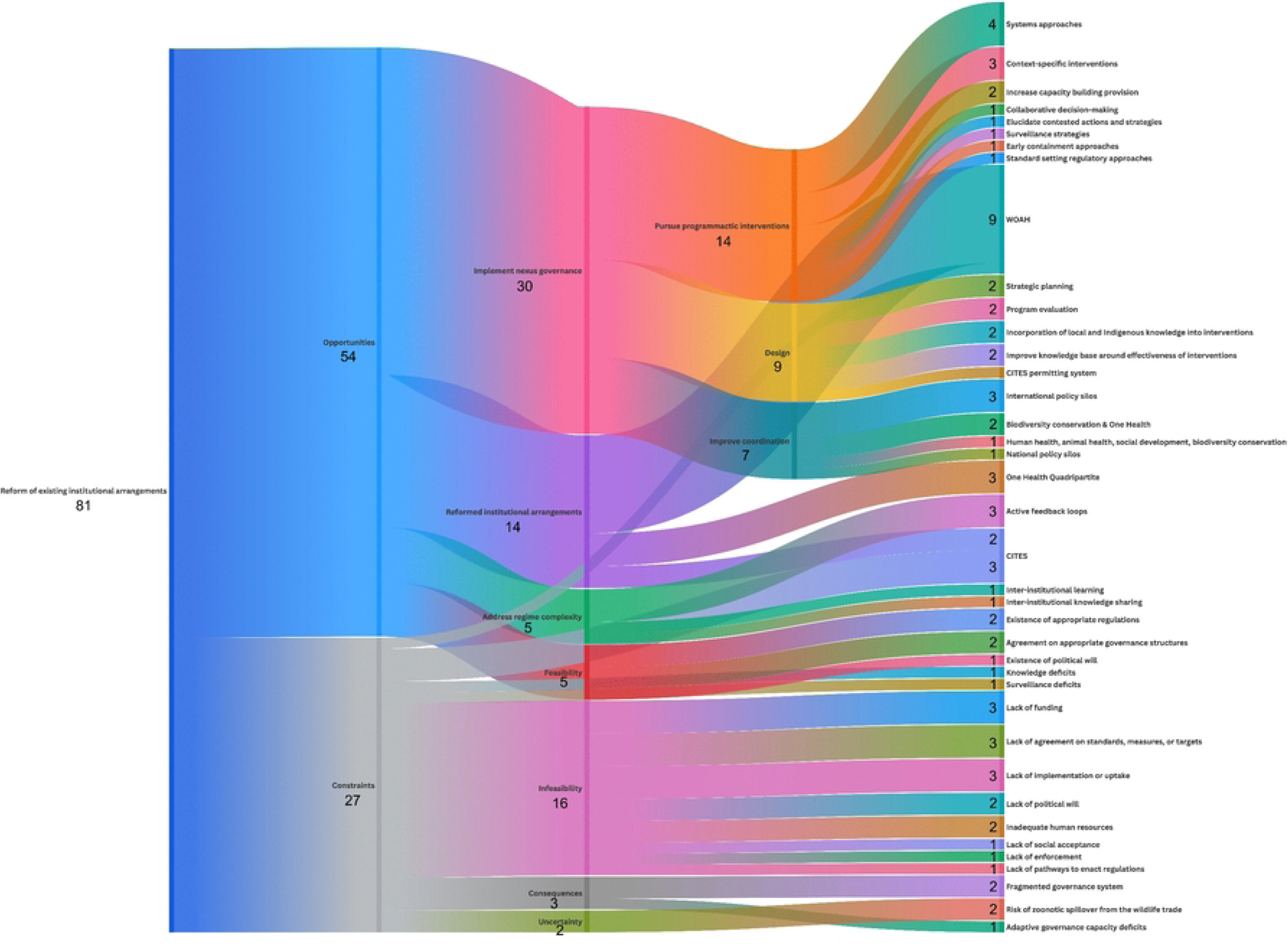
Alluvial diagram for improved use of existing institutional arrangements. Alluvial diagram illustrating flows of opportunities and constraints in response to recommendations for improved use of existing institutional arrangements.

**Fig 6.**
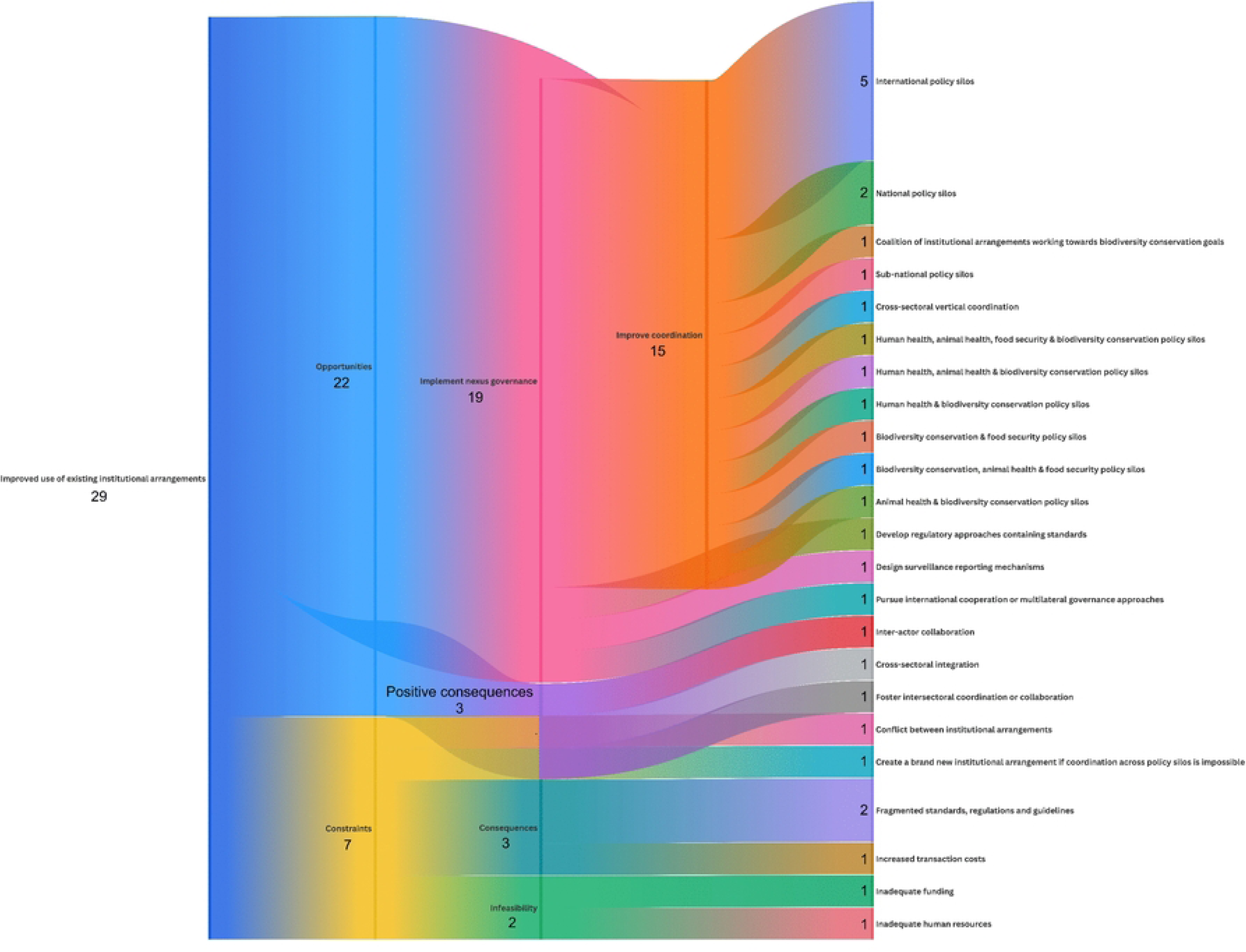
Alluvial diagram for reform of existing institutional arrangements. Alluvial diagram illustrating flows of opportunities and constraints in response to recommendations for reform of existing institutional arrangements. CITES: Convention on International Trade in Endangered Species of Fauna and Flora; WOAH: World Organisation for Animal Health.

**Fig 7.**
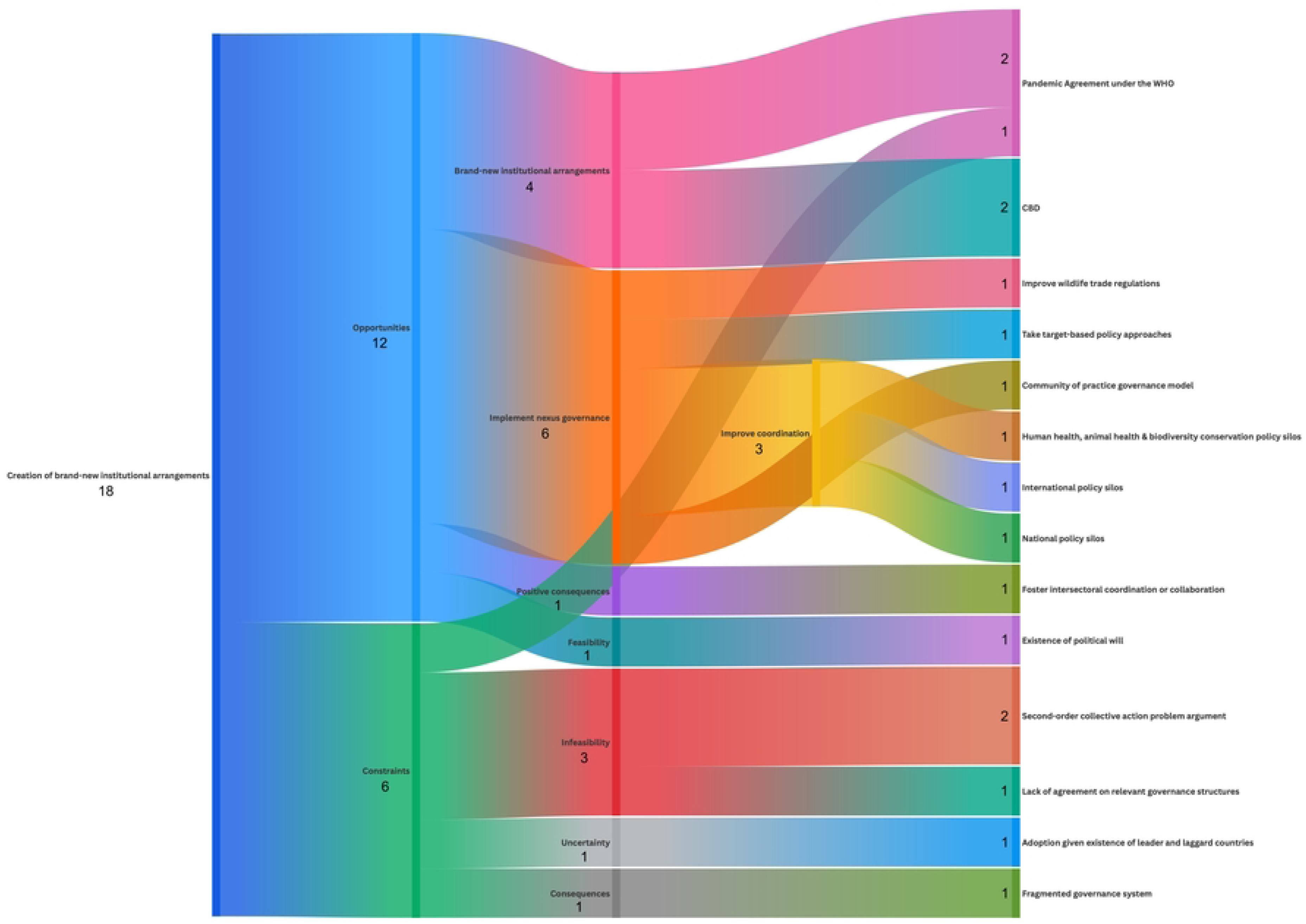
Alluvial diagram for creation of brand-new institutional arrangements. Alluvial diagram illustrating flows of opportunities and constraints in response to recommendations for the creation of brand-new institutional arrangements. CBD: Convention on Biological Diversity; WHO: World Health Organisation.

Diagrams are read from left to right. Each vertical stage denotes the policy position (i.e. leftmost vertical stage), opportunities, constraints, or alternatives (i.e. second from the left vertical stage), theories and overarching themes (i.e. intermediary vertical stages), and specific themes (i.e. rightmost vertical stage). Horizontal flows connect code application values across vertical stages, showing the transition of code application from policy position to various categories to specific recommendations. The relative amount a given variable or recommendation occurred is given at each node or group within a vertical stage.

A network graph was also plotted using data visualisation software *Flourish* [58] to visualise the frequency of the policy sectors KIs recommended improved coordination between to improve the governance of pandemic prevention in relation to the wildlife trade (Fig 8). The nodes in the network analysis are the policy sectors, while the edges represent the relationships or interactions between policy sectors that KIs recommended improved coordination between. The degree centrality of these policy sectors assesses the extent to which policy sectors feature in KI recommendations to improve coordination across policy silos for future governance. Degree centrality was calculated manually using the formula: Degree Centrality (node) = Degree of the node / Maximum possible degree in the network [59] (S9 Table). The node degree for each policy sector was calculated by adding the number of times a policy sector featured in all improved coordination across policy silos code applications. The maximum possible degree in the network was the highest node degree of the policy sectors that featured in the improved coordination across policy silos code applications.

**Fig 8.**
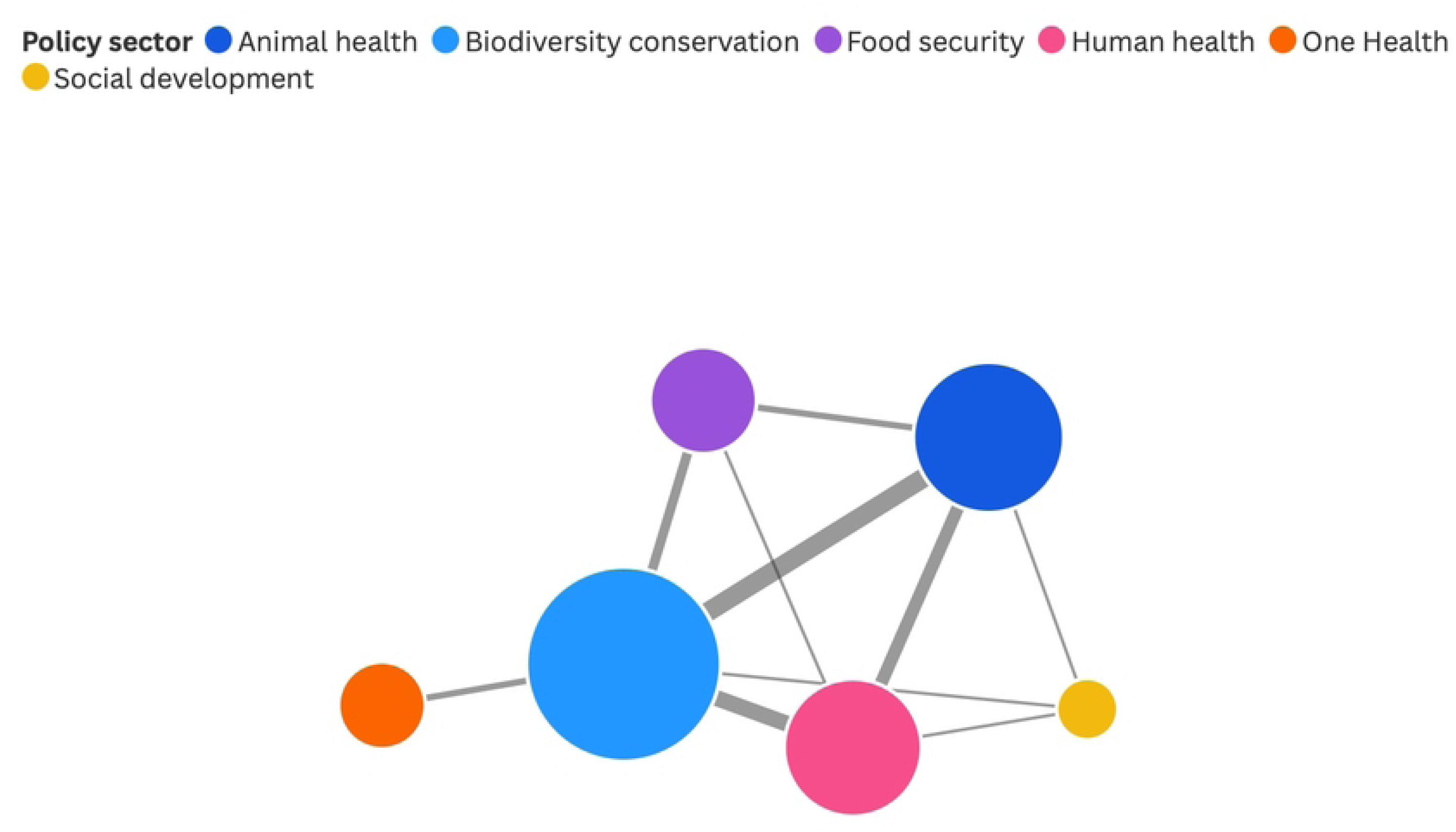
Network graph for improved coordination between siloed policy sectors. Network graph of policy sectors that key informants recommended improved coordination between. The node sizes are a function of which policy sectors most frequently appeared in key informant recommendations for improved coordination, with the largest node representing the most discussed policy sector and the smallest node representing the least discussed policy sector.

## Results

Our results are presented in two main overarching sections: the first section (i. e., overview) presents a summary of the recommendations based on high-level concepts allowing for broad applicability, while the second section (i.e. specific recommendations) presents operationalisable recommendations framed conceptually.

Our dataset contained 52 quotes focused on policy recommendations from a sample of 21 KIs representing six policy sectors (i.e. animal health, biodiversity conservation, food security, human health, international trade, and One Health) and four actor types (i.e. intergovernmental organisation, research institution, treaty secretariat, and nongovernmental organisation). Most recommendations (i.e. 9/13 recommendations = 69%; S10 Table) were raised by more than one KI, representing multiple actor types and policy sectors, while the remaining recommendations were raised by a single KI representing a single actor type and policy sector (i.e. 4/13 recommendations = 31%; S10 Table).

### Overview

Feasible recommendations for improving governance were identified by highlighting opportunities, constraints, and alternatives within the four policy positions as per our research questions. Wildlife trade bans were most often discussed by KIs (i.e. 27/53 code applications = 51%; Fig 2), with livelihood rationales raised as a major constraint against them (i.e. 6/53 code applications = 11.3%; Fig 2). Improved wildlife trade regulations were an ideal alternative to bans (i.e. 5/53 code applications = 9.4%; Fig 2). Reform to existing institutional arrangements were discussed second-most frequently (i.e. 18/53 code applications = 34%; Fig 3), with KIs focusing on the World Organisation for Animal Health (WOAH; formerly the Office International des Epizooties [OIE]) as an institutional arrangement where reform was particularly needed and, importantly, feasible (i.e. 10/53 code applications = 18.9%; Fig 3). KIs less commonly discussed the creation of brand-new institutional arrangements (i.e. 6/53 code applications = 11%; Fig 3), and improved use of existing institutional arrangements (i.e. 2/53 code applications = 4%; Fig 3). KIs raised more constraints than opportunities or alternatives for implementing wildlife trade bans (Fig 2). In contrast, KIs raised more opportunities to realise recommendations than constraints across improved use and reform of existing institutional arrangements and the creation of brand-new institutional arrangements (Fig 3).

Except for recommendations regarding wildlife trade bans (Fig 4), recommendations conformed with the nexus governance (i.e. 68/101 = 67%) and regime complex (i.e. 5/101 = 5%) theoretical approaches (Figs 5-7). Recommendations under nexus governance aimed to achieve policy coherence (i.e. 42/68 opportunities and alternatives = 62%; Figs 5-7) or address sectoral fragmentation (i.e. 26/68 opportunities and alternatives = 38%; Figs 5-7). Sectoral fragmentation was addressed by improving coordination across policy silos and creating a coalition of like-minded actors toward a common goal (Text Box 1). Policy coherence is achieved through addressing knowledge gaps and pursuing various programmatic interventions, governance and policy approaches, and institutional design considerations (Text Box 1). Recommendations aligned with regime complex theory regarding reform of WOAH and involved polycentric knowledge sharing.

Improved coordination between policy sectors featured most prominently across all four policy positions than other recommendations. The biodiversity conservation policy sector featured most often in KI recommendations (i.e. degree centrality = 1; S9 Table), underscoring the need to better integrate the biodiversity conservation policy sector and engage biodiversity conservation actors and institutional arrangements within global pandemic prevention governance (Fig 8). The animal health policy sector featured second most often in KI recommendations for improved coordination between policy sectors (i.e. degree centrality = 0.6; S9 Table). The link between the biodiversity conservation and animal health policy sectors was the strongest of all links between policy sectors, indicating that improved coordination between these two sectors featured most frequently in KI recommendations (S9 Table). The food security (i.e. degree centrality = 0.3), One Health (i.e. degree centrality = 0.2), and social development (i.e. degree centrality = 0.1) policy sectors were mentioned less than average in KI recommendations (i.e. average degree centrality = 0.45; S9 Table), potentially indicating a lower priority from KIs to integrate these sectors in global pandemic prevention governance or that these sectors are already integrated enough or represented within One Health approaches. While integration of multiple policy sectors is inherently One Health in application, One Health is only mentioned within the context of KI recommendations to integrate the biodiversity conservation policy sector within the Quadripartite Collaboration for One Health (Quadripartite hereafter; comprised of the Food and Agricultural Organisation of the United Nations [FAO], the United Nations Environment Programme [UNEP], WHO, and WOAH).

### Specific recommendations

#### Wildlife trade bans

##### Achieve policy coherence through improving understanding of zoonotic spillover causality and effectiveness of interventions

The knowledge base about the causality of zoonotic spillover from the wildlife trade was emphasised as in need of improvement to inform the design of wildlife trade bans and their effectiveness in reducing zoonotic spillover risk from the wildlife trade (KIs #1, #9, & #10). For example, partial bans that prohibit the illegal wildlife trade would not prevent zoonotic spillover risk from the legal wildlife trade (KI #23). Similarly, complete bans on wildlife trade would not prevent zoonotic spillover risk from other causal pathways (e.g. livestock or vector-borne zoonotic diseases; KI #16). Without elucidation of the causality of pandemics emerging from the wildlife trade, a risk exists that the various causal pathways for zoonotic spillover involving different wildlife uses, policy sectors, and actors will further fragment accountability by enabling blame-shifting or making excuses for inaction (KI #16). Similarly, the general support and promotion for a ban on wildlife trade during the onset of the COVID-19 pandemic was partly regarded by KIs as political opportunism for the promotion of an anti-wildlife trade agenda (KI #8). The advancement of political decisions, such as wildlife trade bans, based on the acute onset of the COVID-19 pandemic was viewed as inappropriate, particularly given the uncertainty around the origin of SARS-CoV-2 (KI #8).

##### Achieve policy coherence through improving wildlife trade regulations

Wildlife trade regulations concerning markets and food safety should be improved rather than implement wildlife trade bans (KIs #15 & #18). The supply and demand for wildlife should also be reduced so that wildlife trade value chains are kept to a more manageable size (KIs #14 & #18). For instance, KIs did not agree on whether well-managed wildlife harvests from native ecosystems were less risky than wildlife farming in controlled captive environments (KIs #11, #15, #18), suggesting that improved regulation of the wildlife trade must address knowledge gaps in the causality of zoonotic spillover and account for differences between wild harvests and wildlife farming in reducing zoonotic spillover risk. Improving wildlife trade regulations provides an opportunity to maximise synergies and co-benefits across multiple policy objectives (e.g. create socioeconomic incentives for local populations to engage in sustainable wildlife trade and use, thereby promoting biodiversity conservation and sustainable development; KI #15).

Applying the Hazard Analysis and Critical Control Points approach, which regulates domestic animal trade, to the wildlife trade (KI #10) is an example of improving wildlife trade regulations. This would require the integration of regulations and mechanisms of relevant institutional arrangements for domestic animal trade (e.g. WOAH and FAO) with those for the wildlife trade (e.g. CITES; KI #10).

##### Achieve policy coherence through incorporating human rights within policy responses and ensuring wildlife trade bans are context specific

Wildlife trade bans have the potential to exacerbate trade-offs and unintended consequences. As such, bans were not supported by KIs due to the dependence of Indigenous peoples and local communities on the wildlife trade for livelihoods and food security, the cultural importance of the wildlife trade and hunting for Indigenous and local peoples, the potential for illegal wildlife trade to continue through clandestine economies, and potential co-benefits of fostering community resilience and biodiversity conservation benefits from a sustainable legal wildlife trade (KIs #1, #9, #10, #12, #15, #16, & #19). Enacting bans without consideration of the drivers of demand would drive the wildlife trade to underground economies or clandestine markets where the trade will continue illegally without regulation or supervision (KIs #1, #9, & #10).

Consequently, surveillance of potential disease risk would be made more complicated and untraceable and would negatively impact the ability to assess and mitigate zoonotic spillover risk (KI #18). Furthermore, wildlife trade bans would prioritise biodiversity conservation and pandemic prevention priorities over fostering community resilience to climate change through providing diversified, climate-resilient income and livelihood streams and creating incentives for biodiversity conservation locally (KIs #12 & #15).

Applying blanket wildlife trade bans would be inappropriate and ineffective given sociocultural issues, pathogens, and species that vary between jurisdictions (KIs #12 & #18), and if wildlife trade bans do not consider and address the drivers of demand for wildlife (KI #18). Furthermore, enacting bans would be ineffective and infeasible in jurisdictions with law enforcement deficits to ensure compliance with existing, or future, wildlife trade regulations (KI #12). For example, cases where wildlife trade and farming are done safely according to relevant standards and regulations, such as in South Africa and Botswana, challenge narratives of bans as a panacea to prevention of pandemics emerging from zoonotic spillover (KI #16).

Given the potential for trade-offs and unintended consequences, policy responses for zoonotic spillover prevention must incorporate human rights approaches in designing and implementing interventions that regulate the wildlife trade (KIs #1, #9, #12 #15, #16, & #19). Implementation of wildlife trade bans should also consider the sociopolitical, cultural, economic, and ecological contexts (e.g. species, wildlife products, geography; KI #18) and monitor for unintended consequences (KI #1).

#### Improved use of existing institutional arrangements

##### Achieve policy coherence through harmonising siloed international regulatory systems

Integrating the development of future standards, regulations, or guidelines for pandemic prevention in relation to the wildlife trade within the work of existing relevant institutional arrangements was advised as a strategy to reduce fragmentation and conflict between intersecting global governance systems that comprise global pandemic prevention, address gaps in the governance system, and achieve regulatory harmonisation (KI #7). For example, future standards for the trade of wildlife for food could be integrated in the work of the existing Codex Alimentarius Commission, as the Commission is mandated with food safety and the food trade (KI #7).

##### Achieve policy coherence through enhancing recognition of existing collaborations and fora

Enhancing awareness of existing collaborations would support more effective use of current institutional arrangements than creating new ones (KI #1). For example, collaboration and coordination between WOAH, the International Union for Conservation of Nature (IUCN), CITES, the Convention on Biological Diversity (CBD), and the Wildlife Conservation Society (WCS) creates a coalition that mobilises the animal health, biodiversity conservation, and international trade policy sectors towards primarily biodiversity conservation ends (KI #1).

Optimising the use of existing collaborations and fora would address the second-order collective action problem of new institutional arrangements diverting political will and human and financial resources from ongoing technical progress made at the working level (KI #1) by leveraging existing mechanisms and resources for enforcement, compliance, and coordination.

Similarly, existing institutional arrangements should be better organised to address gaps in global governance for pandemic prevention through the United Nations (UN; KI #16). The perceived neutrality of the UN as a democratic decision-making body enables the equal participation by member states and ensures their engagement by not serving the interests of individual member states (KI #16).

##### Address sectoral fragmentation through improving coordination between siloed policy sectors

Achieving cross-sectoral horizontal integration through improved coordination between the animal health, biodiversity conservation, food security, and human health policy sectors was emphasised (KIs #6, #8, #9, #15, & #16). Specific mechanisms were mentioned as enabling coordination between these policy sectors:

- The 2015 Cooperation Agreement between WOAH and CITES is a mechanism for improved collaboration between the animal health and biodiversity conservation policy sectors as this Agreement defines specific objectives and opportunities to collaborate and complement each other. Subsequent efforts included the release of the 2021 Wildlife Health Framework [60] and the ongoing renewal of the 2015 Cooperation Agreement (at the time the interview was conducted), and more recently through a 2024 Memorandum of Understanding to address wildlife health regulation gaps between WOAH and CITES (KI #6; [61]).
- The CBD joining a 2021 Expert Working Group on Biodiversity, Climate, One Health and Nature-Based Solutions between the WHO and the IUCN would create an opportunity to further improve coordination between the human health and biodiversity conservation policy sectors (KI #9). This Expert Working Group leverages progress made towards cross-sectoral integration by the 2015-2020 Interagency Liaison Group on biodiversity conservation and health, cochaired by the CBD and WHO.

In addition to these existing mechanisms, KIs recommended the development of new cross-sectoral mechanisms:

- A working group comprised of UNEP, the Convention on the Conservation of Migratory Species of Wild Animals (CMS), CBD, and FAO may improve coordination between the biodiversity conservation and food security policy sectors (KI #16). Such a working group would consider wildlife from a biodiversity conservation management standpoint and focus on the impact of disease on wildlife populations given the institutional arrangements’ relevant technical expertise and programming (KI #16). However, limited funding and human resources prevents the creation of this working group (KI #16).
- Increasing collaboration between CITES, FAO, WOAH, and other institutional arrangements mandated with animal health and veterinary conditions may improve coordination between the biodiversity conservation, food security, and animal health policy sectors at the international level (KI #15). Were coordination across these policy sectors infeasible, the creation of a new institutional arrangement to mainstream biodiversity conservation governance would be a suboptimal alternative as a new institutional arrangement would overlap with the mandates of these institutional arrangements (KI #15).
- A joint committee between WHO, WOAH, and CITES may improve intersectoral collaboration between the human health, animal health, and biodiversity conservation policy sectors (KI #8). In such a joint committee, the constituent institutional arrangements would report the findings and rulings of the joint committee to their respective governing bodies (KI #8).
- The creation of comprehensive surveillance databases is recommended to integrate and address existing gaps between the surveillance systems of the human and animal health policy sectors (i.e. WOAH’s World Animal Health Information System and the surveillance system of the WHO’s IHR), and to improve coordination and information sharing between these policy sectors (KI #9).

A global governance practitioner KI emphasised that national and sub-national government departments, ministries, and policy sectors do not collaborate or coordinate with each other. This is demonstrated by policies that conflict with each other and competition for funding and political will through government agenda and priority setting. To promote a cross-sectoral One Health approach, horizontal coordination across the human health, animal health, food security, and environmental health policy sectors that occurs internationally through the Quadripartite must be similarly realised at the national and sub-national levels (KI #7) through structural reform that enables shared governance approaches.

#### Reform of existing institutional arrangements

##### Address sectoral fragmentation through expanding Quadripartite membership to include the CBD and CITES

The addition of UNEP into the Quadripartite collaboration in 2022 demonstrates the political will to incorporate relevant policy sectors into the operationalisation of One Health. This provides precedent for expanding the membership of the Quadripartite to include additional institutions with overlapping, and interdependent, mandates. Cross-sectoral horizontal integration across the biodiversity conservation and One Health policy sectors at the international level may be improved by adding CITES (KI #6) and the CBD (KI #16) to the Quadripartite. Similarly, a member of the Quadripartite is recommended to take over the International Alliance Against Health Risks in Wildlife Trade, a collaborative platform that aims to reduce zoonotic spillover risk (KI #19). It is unclear whether such a reform would impact the Quadripartite as an institutional arrangement (e.g. the Alliance would become a new member of the Quadripartite) nor how a Quadripartite member would subsume the Alliance (e.g. merger, acquisition, or an agreement to collaborate).

##### Achieve policy coherence through expanding WOAH’s mandate to include the wildlife trade

WOAH’s mandate could be expanded to regulate the wildlife trade in relation to pandemic prevention given recognition by the WTO’s SPS Agreement as the reference organisation for providing standards on zoonoses and terrestrial and aquatic animal health in international trade (KIs #3-5). The development of new institutional arrangements, other than those by WOAH, would not automatically be recognised by the SPS Committee and would require consensus within the Committee to recognise the resulting new institutional arrangements, potentially exacerbating fragmentation and policy incoherence. This recognition of WOAH as a reference organisation demonstrates agreement on the relevant governance structures and the existence of feasible pathways to enact needed changes. However, implementing reforms advised by WOAH may not be feasible for low- and middle-income member states due to limited funding and human resources available for programs related to the wildlife trade (KI #12). As such, expanding WOAH’s mandate should be coupled with strengthening its capacity to provide guidance and capacity building that is adaptive and specific to member state circumstances (KI #12).

##### Address regime complexity through leveraging active feedback loops to enhance institutional reflexivity, inter-institutional learning, and knowledge management

Active feedback loops were included in recommendations of WOAH reform, with mechanisms often taking the form of working groups or consensus processes that enable implementation evaluation, analysis, knowledge dissemination, and iterative updating of agenda setting and policy formulation [57]. As working groups or consensus processes involve the development of networks of stakeholders, experts, and beneficiaries, these mechanisms may improve coordination and collaboration between institutional arrangements from different regimes to facilitate information sharing and joint knowledge production. For example, WOAH may collaborate with the Cochrane Collaboration or the International Initiative for Impact Evaluation to inform the development of a new Collaborating Centre for Evidence-Based Animal Health Policy to complement and support implementation of guidance to member states [57]. While only recommended within the context of WOAH reform, working group mechanisms can be applied to improving wildlife trade regulations, improving the use of or reforming existing institutional arrangements, and creating brand-new institutional arrangements to promote active feedback loops, inter-institutional learning, and knowledge management.

##### Achieve policy coherence through expanding CITES’ mandate to consider pandemic risk

Feasible potential options for reforming CITES includes incorporating animal health issues within its remit, banning live animal trade in CITES listed species, regulating domestic ‘wet’ markets, and improving collaboration with WOAH and other institutional arrangements (KI #15). Adding animal health and veterinary health conditions to the CITES permitting system, such as including veterinary controls, transportation practices, and disease-free statuses, would be feasible given the flexibility for member states to set their own standards (KI #15). However, these reformed compliance mechanisms would only apply to species listed within CITES, based on conservation concerns. This change would dilute the already limited financial resources available to implement CITES’ biodiversity conservation agenda; require technical expertise, funding, and human resources that CITES currently lacks; involve complex multilateral treaty negotiations; and provide an opportunity for member states to weaken CITES’ enforcement capability (KI #15). Such negotiations are therefore unlikely to occur in the short term and may instead occur 10-15 years in the future. The existence of two regulatory systems for signatory and non-signatory parties risks exacerbating fragmentation and policy incoherence through using CITES for pandemic prevention aims and the potential for contradictory or competing claims during international trade between signatory and non-signatory parties (KI #15). Similarly, there is limited political will among CITES member states to agree upon and implement a list of animal species with potential risk for zoonotic spillover, including consideration for banning or strictly regulating the trade of these species (KI #15).

#### Creation of brand-new institutional arrangements

##### Achieve policy coherence through establishing an intergovernmental panel on pandemic risk and evidence

The creation of a coordinating mechanism that fosters intersectoral coordination is preferred to a brand new permanent institutional arrangement, as permanent institutional arrangements require additional resources for administration that could instead be used for technical, programmatic work (i.e. second-order collective action problem; KI #12). Considering this rationale, the establishment of an intergovernmental evidence panel on pandemic risk was proposed (KI #12), similar to the Intergovernmental Panel on Climate Change [62], the Independent Panel for Evidence for Action Against Antimicrobial Resistance [63], or an Intergovernmental Panel for

One Health to strengthen pandemic prevention, preparedness, and response [64]. By having a mandate to integrate actors and share knowledge rather than focus on a singular issue, such coordinating mechanisms enable greater flexibility to address multiple issues (KI #12). This type of intergovernmental panel, known as science-policy platforms, would adopt a community of practice model, where expertise and local community participation can collaborate based on a common concern, allowing for greater inclusivity and participation (KI #12).

##### Achieve policy coherence through developing an anticipated Pandemic Agreement under the WHO

As interviews were undertaken prior to formal discussions around the Pandemic Agreement, KIs proposed that this type of agreement developed under the WHO could address sectoral fragmentation by improving cross-sectoral coordination across the human health, animal health, and biodiversity conservation policy sectors at the international and national levels (KI #23). The Agreement would address the paucity of multilateral guidance available at the time by providing a framework related to pandemic prevention and supporting capacity strengthening in member states (KI #23).

However, KIs proposed that the development of a Pandemic Agreement by the WHO may be constrained by concerns about institutional niche. Such an agreement would intersect with WOAH’s area of competence and practice as it would involve animal diseases and veterinary services (KI #15). This overlap and potential for tension between WHO and WOAH demonstrates a lack of agreement regarding the governance structures involved in regulating the wildlife trade in relation to pandemic prevention. Furthermore, KIs expressed concern that the adoption of a Pandemic Agreement could be constrained by a lack of consensus and political will among member states and the second-order collective action problem of enforcement and monitoring state compliance (KI #23).

##### Achieve policy coherence through developing a new target-based framework under the CBD

The CBD could take a target-based approach by creating a new framework with targets and indicators that parties can commit to, implement, and deliver on (KI #23). This recommendation is based on political will for the CBD’s biodiversity conservation targets, demonstrated by the UN General Assembly endorsing the 2010 Aichi Biodiversity Targets and the goals and targets of the 2024 Kunming-Montreal Global Biodiversity Framework, which replaced and updated the Aichi Targets (KI #23). The drafting and negotiation of the CBD’s Global Action Plan on Biodiversity and Health provided an opportunity to improve wildlife trade regulations by incorporating guidance and best practice that relates biodiversity conservation to zoonotic spillover risk, such as not trading live animals, enacting sanitary measures at markets, and butchering meat away from markets, *inter alia* (KI #23).

## Discussion and conclusion

We found that international governance entrepreneurs and practitioners engaged in pandemic prevention perceive reform of WOAH and the breaking down of policy silos as the primary way to improve future governance for pandemic prevention in relation to the wildlife trade for human consumption. As WOAH is under-resourced as a non-UN organisation, enhanced funding would be imperative to enable a strengthened and expanded mandate. Enabling shared governance within the global regime complex between WHO whose leadership oversaw the pandemic treaty, to more comprehensively encompass and address animal and environmental health concerns as upstream sources of emergent zoonosis through relevant multilateral organisations would be imperative for governance of pandemic prevention.

Notably, there was wide concern for how the creation of a brand-new institutional arrangement tasked with pandemic prevention could potentially exacerbate existing governance fragmentation and potentially leave governance deficits unaddressed. However, implementing the recommendations for improved use and reform of existing institutional arrangements is challenged by power dynamics, insufficient human resources, limited funding, and enforcement deficits within the global governance regime complex.

The recommendations captured and analysed here demonstrate how a nexus governance approach is central to advancing pandemic prevention in light of these challenges and trade-offs within the global governance regime complex. Panaceas of synergies across policy sectors need to be considered with cautious optimism and scrutinised given the potential for unintended consequences to arise. Although wildlife trade bans are unlikely to effectively advance pandemic prevention, balanced and context-specific wildlife trade regulations promise to help advance this agenda alongside animal health, biodiversity conservation, and food security objectives.

As the WHO’s Pandemic Agreement negotiations largely focused on pandemic preparedness and response with only general statements on One Health prevention to address ecological and animal drivers of zoonotic spillover through a coordinated multisectoral stance (Articles 4 and 5 [10]), there remains potential to mainstream consideration of specific approaches to more fully address upstream prevention of zoonotic spillover from the wildlife trade at both global and national governance levels. The recommendations from our analysis address governance gaps through promoting shared intersectoral governance approaches [65], which complement and support recent governance innovations [66]. In the wake of the COVID-19 pandemic, an active literature strand on pandemic prevention recommendations rapidly emerged. Our findings confirm that a renewed, theoretically guided literature strand on the topic is warranted in the wake of the governance gaps left by the Pandemic Agreement, which could be informed by the Earth System Governance research framework [67].

## Data Availability

The data that support the findings of this study are available on request from the corresponding author. The data are not publicly available due to privacy or ethical restrictions.

## Acknowledgements

The authors would like to thank the key informants who were interviewed as part of this study for generously contributing their time and insights and Drs. Chloe Clifford Astbury and Eva-Maria Nag for their valuable comments on previous drafts.

## Supporting information

**S1 Text. List of interview guide questions from which data was collected.**

**S2 Table. Codebook used in the thematic analysis of quotes.**

**S3 Text. Results from publicly available documents referred to by key informants during interviews.**

**S4 Table. Frequency of discourse code application.**

**S5 Table. Flow of wildlife trade ban code applications to rationales against bans, rationales for bans, alternatives to bans, theories, and overarching and specific themes.**

**S6 Table. Flow of improved use of existing institutional arrangement code applications to opportunities, constraints, theories, and overarching and specific themes.**

**S7 Table. Flow of reform of existing institutional arrangement code applications to opportunities, constraints, theories, and overarching and specific themes.**

**S8 Table. Flow of creation of brand-new institutional arrangement code applications to opportunities, constraints, theories, and overarching and specific themes.**

**S9 Table. Policy sectors by number of occurrences in all code applications, connections between policy sectors, and degree centrality calculations.**

**S10 Table. Recommendations by the specific actor type and policy sector and the proportion of actor type, policy sector, and key informants that raised a recommendation.**

**S11 Table. Examples of codes applied to quotes identified from interviews with key informants.**

**S12 Table. Overview of recommendations for potential real-world governance responses for pandemic prevention from wildlife trade for human consumption.**

